# Association between Serum atherogenic index and cardiovascular diseases and mortality in early adulthood (18-44 years old):Kailuan Longitudinal Cohort Study

**DOI:** 10.1101/2025.10.10.25337777

**Authors:** Mianwang He, Nana Yin, Chi Wang, Zekun Feng, Shouling Wu, Hao Xue

**Affiliations:** Department of Neurology, First medical center, Chinese PLA General hospital, Chinese PLA medical school, Beijing 100853, China; Department of Cardiology, Second Medical Center, Chinese PLA General Hospital, Chinese PLA Medical School, Beijing 100853, China; Department of Cardiology, Sixth Medical Center, Chinese PLA General Hospital, Chinese PLA Medical School, Beijing 100853, China; Department of Cardiology, Kailuan General Hospital, Tangshan 063000, China

**Keywords:** Atherogenic index of plasma, Cardiovascular diseases, Stroke, All-cause mortality, Early adulthood

## Abstract

**BACKGROUND:** The atherogenic index of plasma (AIP), calculated as log (triglycerides / high-density lipoprotein cholesterol), has emerged as a novel marker for assessing atherogenic risk and cardiometabolic health. However, the relationship between AIP and the risks of cardiovascular diseases (CVD) and all-cause mortality in early adulthood remains unclear. In the present study, we investigated the association of AIP with CVD and all-cause mortality in a large-scale cohort.

**METHODS:** A total of 41,828 participants aged 18 to 44 years without pre-existing CVD at baseline were enrolled from surveys during 2006 to 2016, and were categorized according to AIP quartiles. All participants were followed biennially through December 31, 2022. To assess the associations between AIP and the incidence of CVD, stroke, and all-cause mortality, both univariate and multivariate Cox proportional hazards regression models were used. Additionally, Kaplan–Meier analysis was conducted to compare cumulative incidence of CVD and stroke across AIP quartiles.

**RESULTS:** During an average follow-up of 12.65±3.59 years, a total of 1,113 cases of CVD, 953 cases of stroke, and 969 cases of all-cause mortality were identified. After adjustment for conventional cardiovascular risk factors, including age, gender, smoking status, alcohol consumption, heart rate, hypertension history, triglycerides, total cholesterol, fasting blood glucose, and estimated glomerular filtration rate, the multivariate-adjusted hazard ratios (HRs) and their 95% confidence intervals (CIs) for CVD and stroke, compared to the first quartile (Q1), were as follows: 1.21 (0.99–1.48) and 1.16 (0.94–1.43) in the second quartile group (Q2); 1.44 (1.19–1.74) and 1.28 (1.05–1.58) in the third quartile group (Q3); and 1.40 (1.12–1.75) and 1.28 (1.01–1.63) in the fourth quartile group (Q4). No significant association was observed between AIP and all-cause mortality.

**Conclusions:** Our study found that an elevated AIP is associated with an increased risk of CVD and stroke in young adults. These findings highlight that AIP may serve as a potential risk stratification factor in young populations.

## Introduction

Cardiovascular diseases (CVD), primarily comprising stroke and ischemic heart disease, remain the leading cause of global mortality, accounting for approximately one-third of all deaths worldwide^1^. Although significant progress has been made in assessing traditional risk factors, residual risk has driven the exploration of novel biomarkers^2–5^. The atherogenic index of plasma (AIP), calculated as the logarithmic ratio of triglycerides to high-density lipoprotein cholesterol, is considered a promising indicator of atherogenic dyslipidemia and cardiovascular risk ^6^and is correlated with diabetes mellitus, hypertension, and metabolic syndrome^7–9^. Moreover, AIP reflects pathophysiological disturbances in lipid metabolism^10,11^, thereby offering insights beyond conventional lipid parameters. While previous studies on the association between AIP and CVD were mainly based on populations at high risk, such as individuals with diabetes ^12–14^and non-diabetic hypertensive elderly^15^, evidence in the general population remains limited^16,17^, and is particularly scarce in population of early adulthood. Furthermore, given the increasing recognition that early exposure to cardiovascular risk factors significantly impacts later pathogenesis of CVD^18–21^, the identification of young adults at risk is crucial. Although clinical manifestations of CVD typically emerge after age 45^21^, the atherosclerotic process begins decades earlier. In fact, lipid monitoring before age 40 identifies a majority of individuals with a high likelihood of lifetime elevated lipid levels who consequently have a high long-term risk for CVD^22^. Therefore, we investigated whether an elevated AIP before age 45 independently predicts future CVD, stroke, and all-cause mortality.

## Methods

### Study Participants

The Kailuan Study is an ongoing prospective cohort study based in Tangshan, China. Conducted in accordance with the Declaration of Helsinki, the study protocol was approved by the Ethics Committee of Kailuan General Hospital. The cohort consists of employees and retirees of the Kailuan Group, a coal mining company located in Tangshan. Biennial health examination including all cohort participants were conducted since 2006. All study participants provided their informed consents. Detailed descriptions of the study design and procedures have been published previously^23,24^.

For this analysis, we initially included 47,125 adults aged 18 to 44 years who had undergone baseline health examinations between 2006 and 2016. After excluding 889 participants with pre-existing cardiovascular diseases or use of lipid-lowering medications, 3,514 with missing AIP data, and 894 with incomplete blood pressure measurements, a total of 41,828 participants were eligible for the current analysis. Figure 1 illustrates the participant selection process and study design.

**Fig. 1.**
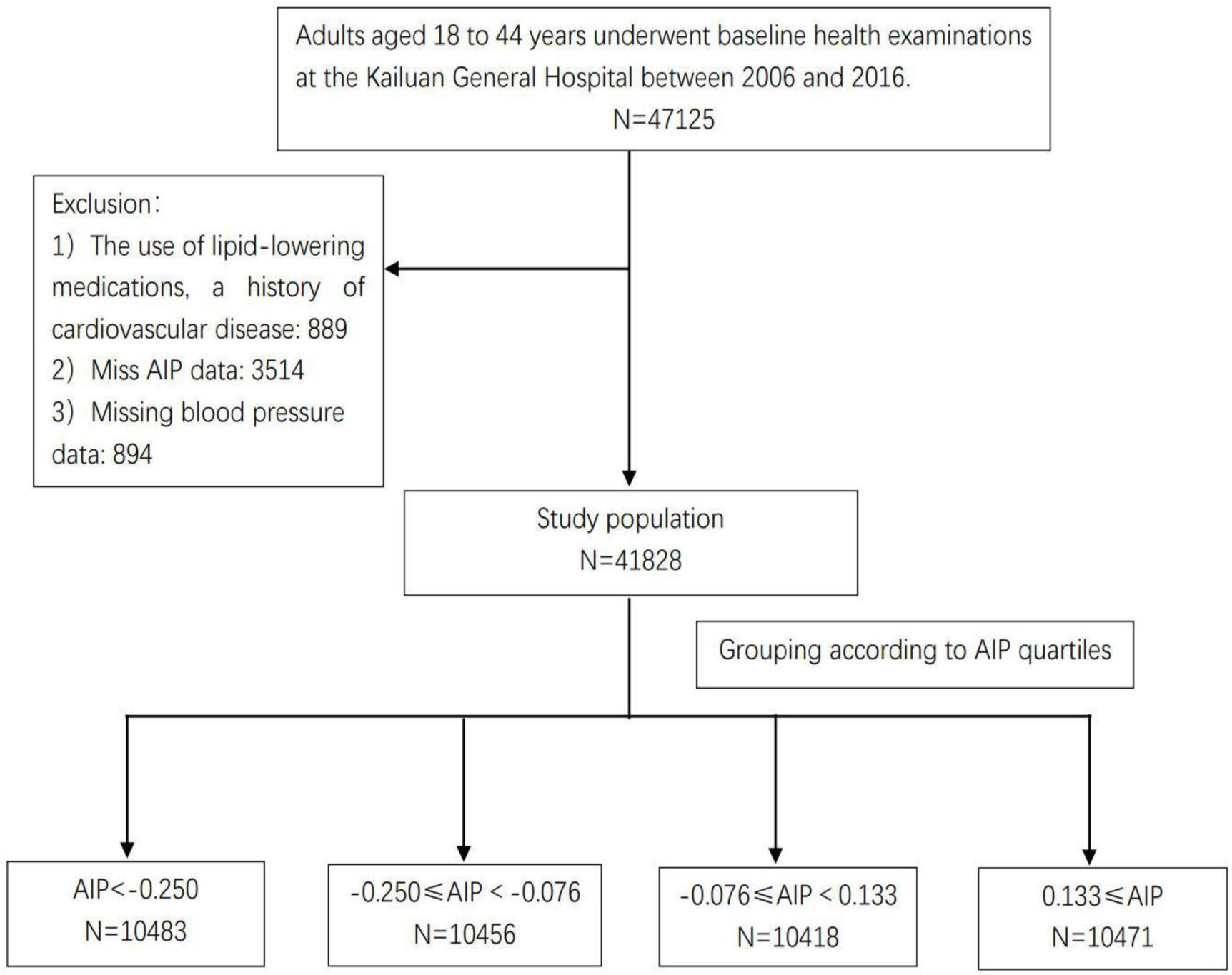
Study population enrollment. AIP: atherogenic index of plasma

### Measurements of AIP and Other Covariates

Baseline data were collected using standardized questionnaires, anthropometric measurements, and laboratory blood tests. All participants provided fasting blood samples after an overnight fast during each physical examination. Blood samples were analyzed on the same day using an auto-analyzer (Hitachi 747; Hitachi, Tokyo, Japan). The biochemical parameters assessed included fasting blood glucose (FBG), triglycerides (TG), total cholesterol (TC), low-density lipoprotein cholesterol (LDL-C), high-density lipoprotein cholesterol (HDL-C), and serum creatinine.

The atherogenic index of plasma (AIP) was calculated as log₁₀[triglycerides (mmol/L) / HDL-C (mmol/L)]. Body mass index (BMI) was computed as weight in kilograms divided by the square of height in meters. Participants were classified as ever-smokers if they had a history of smoking or were current smokers, and as ever-drinkers if they had a history of alcohol consumption or were current drinkers. The estimated glomerular filtration rate (eGFR) was calculated using the Chronic Kidney Disease Epidemiology Collaboration (CKD-EPI) creatinine equation(eGFR) ^25^. Hypertension was defined as systolic blood pressure ≥ 140 mmHg or diastolic blood pressure ≥ 90 mmHg, use of antihypertensive drugs, or self-reported history of physician-diagnosed hypertension. Diabetes was defined as fasting blood glucose ≥ 126 mg/dL (7.0 mmol/L), or treatment with antidiabetic drugs, or self-reported physician-diagnosed type 2 diabetes.

### Definition of outcomes

The primary outcomes of this study were the first occurrence of CVD, stroke, and all-cause mortality. CVD cases included myocardial infarction (MI), ischemic stroke (IS), and hemorrhagic stroke (HS). These events were identified using the following ICD-10 codes: I21 for MI, I63 for IS, and I60–I61 for HS. Data on CVD diagnoses were obtained from the Municipal Social Insurance Institution and the Hospital Discharge Register and were updated annually throughout the follow-up period. An expert panel collected and reviewed annual discharge records from 11 local hospitals to identify patients with suspected CVD. Myocardial infarction was diagnosed according to the World Health Organization’s Multinational Monitoring of Trends and Determinants in Cardiovascular diseases (MONICA) criteria^26^, based on clinical symptoms, electrocardiographic findings, and dynamic changes in myocardial enzyme levels. Stroke was diagnosed in accordance with World Health Organization criteria^27^, involving neurological signs, clinical symptoms, and neuroimaging results—such as computed tomography or magnetic resonance imaging. All-cause mortality data were collected from provincial vital statistics offices and reviewed by physicians.

### Statistical analysis

Participants were stratified into quartiles (Q1–Q4) based on their AIP values. Continuous variables were compared using analysis of variance (ANOVA) or the Kruskal–Wallis test, as appropriate for their distribution, and categorical variables were compared using the chi-square test.

Incidence rates of CVD, stroke, and all-cause mortality were calculated per 1,000 person-years across AIP quartiles. We used Cox regression ^28^ to calculate hazard ratios (HRs) with 95% confidence intervals (CIs) for CVD, stroke, and all-cause mortality among participants in higher AIP quartiles (Q2–Q4), using the lowest quartile (Q1) as the reference group.

In the analysis, Model 1 was unadjusted, Model 2 was adjusted for age and gender, and Model 3 included adjustments for a broader set of variables, including age, gender, smoking status, alcohol consumption, heart rate, hypertension history, triglycerides, total cholesterol, fasting blood glucose, and eGFR. Kaplan–Meier analysis was performed to compare cumulative incidence of CVD and stroke across AIP quartiles. Additionally, we calculated HRs for specific CVD subtypes, including MI, IS, and HS.

To test the robustness of our findings, we further conducted three sensitivity analyses. First, we excluded outcome events occurring within first 2 years of the follow-up period to minimize potential reverse causation. Second, to avoid the influence of cancer history on our results, we excluded participants with cancer at baseline. Third, to minimize the influence of treatment on our results, we excluded users of antihypertensive, hypoglycemic, or lipid-lowering drugs. Finally, we conducted stratified analyses of the associations between AIP and CVD, stroke, and all-cause mortality by sex (men/women), BMI categories (<24 kg/m² vs ≥24 kg/m²), Fasting blood glucose (<7 mmol/L vs ≥7 mmol/L), and Hypertension status (yes/no), respectively.

All analyses were performed using SAS version 9.4 (SAS Institute, Cary, North Carolina) and R software version 3.6.0 (R Core Team, Vienna, Austria). All statistical tests were 2-sided, and p < 0.05 was considered statistically significant.

## Results

### Baseline Characteristics

A total of 41828 participants were included in this study, with an average age of 37.08±6.46 years (33675 men [80.5%] and 8153 women [19.5%]). According to the AIP based on quartile, participants were stratified into quartiles: Q1 (AIP < –0.250; n = 10483), Q2 (–0.250 ≤ AIP < –0.076; n = 10456), Q3 (–0.076 ≤ AIP < 0.133; n = 10418), and Q4 (AIP ≥ 0.133; n = 10471).Baseline characteristics across AIP quartiles are summarized in Table 1. We observed that the following parameters all increased significantly across ascending AIP quartiles: age; levels of FBG, TG, TC, and LDL-C; heart rate; waist circumference; hip circumference; waist-to-hip ratio; BMI; systolic and diastolic blood pressure; the proportion of males; and the prevalence of hypertension and diabetes. In contrast, eGFR, HDL-C levels, and the proportions of nonsmokers and nondrinkers progressively decreased with higher AIP quartiles.

**Table 1.**
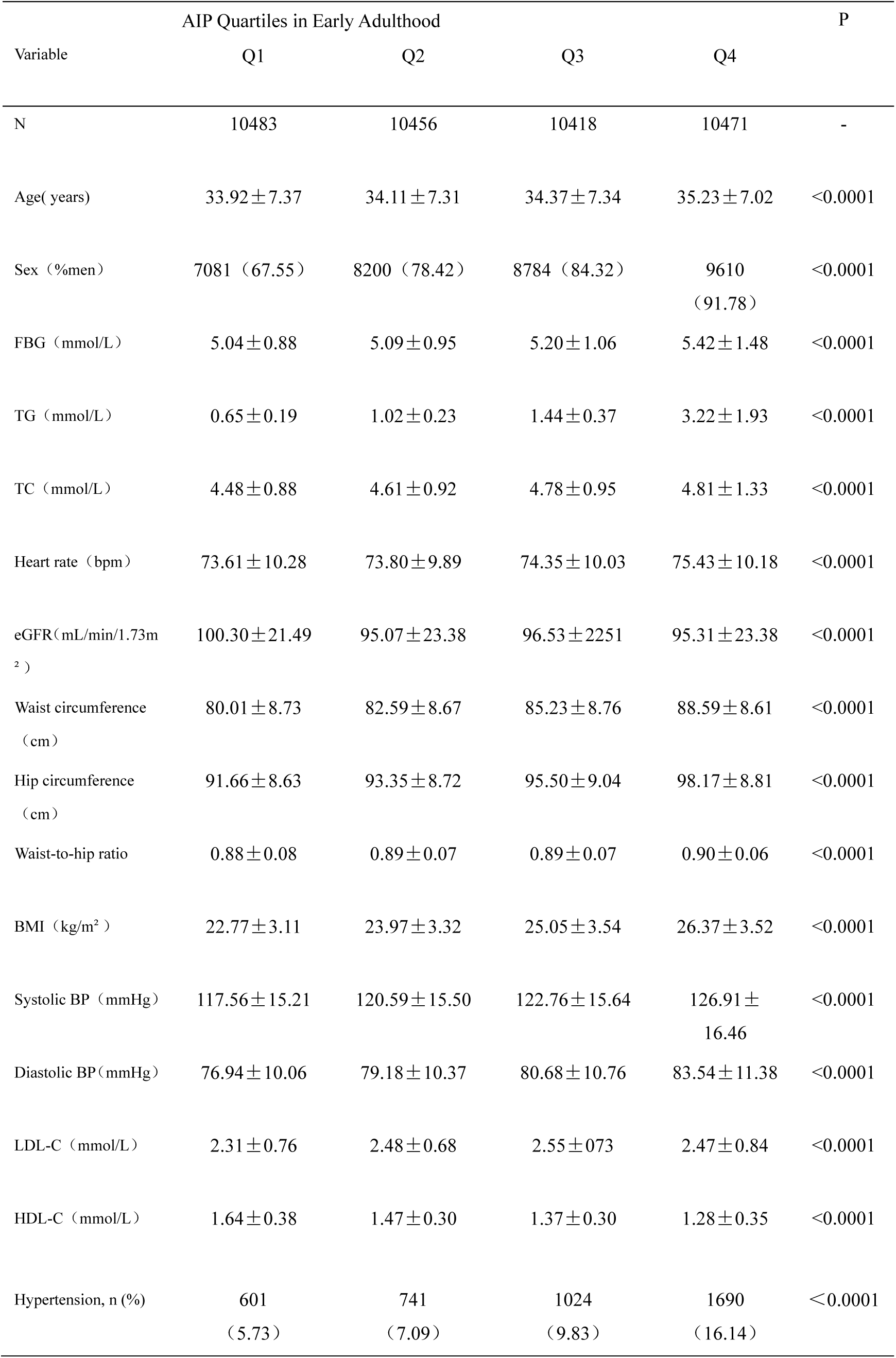
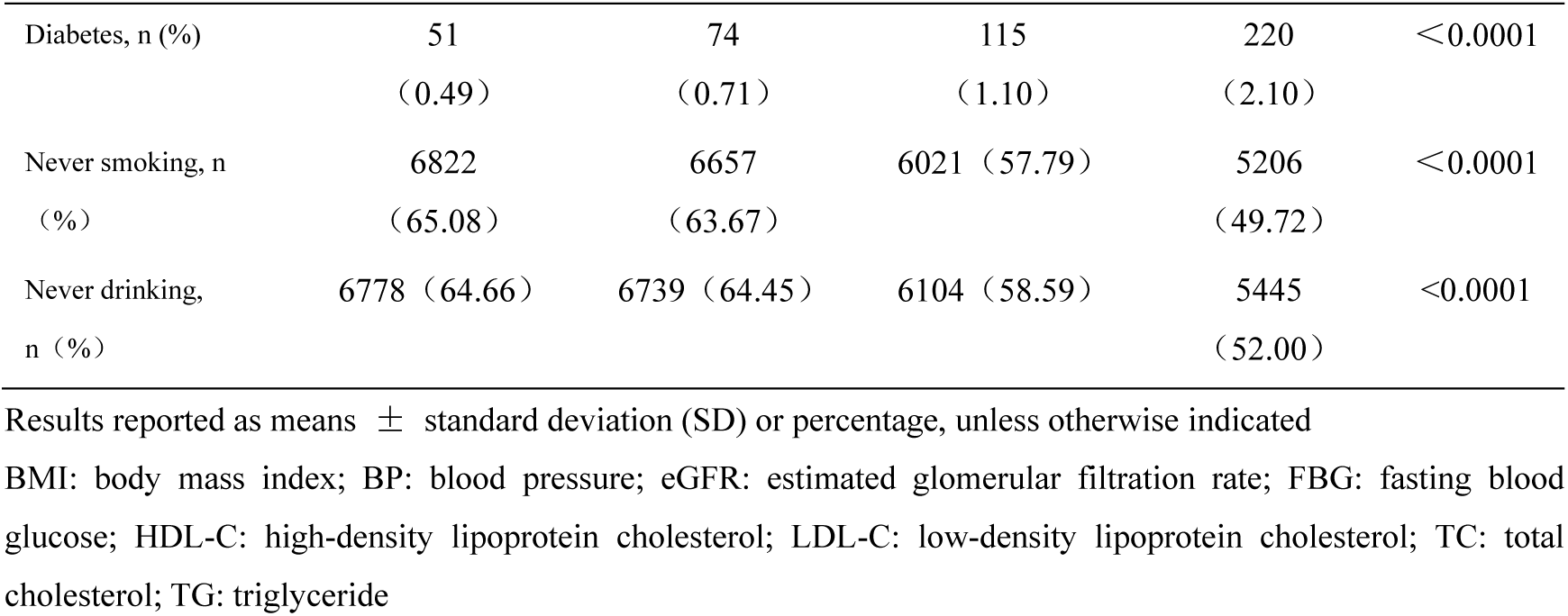
Baseline Characteristics of Participants According to AIP Quartiles in Early Adulthood.

| Variable | AIP Quartiles in Early Adulthood |  |  |  | P |
| --- | --- | --- | --- | --- | --- |
|  | Q1 | Q2 | Q3 | Q4 |  |
| N | 10483 | 10456 | 10418 | 10471 | - |
| Age( years) | 33.92±7.37 | 34.11±7.31 | 34.37±7.34 | 35.23±7.02 | <0.0001 |
| Sex (%men) | 7081 (67.55) | 8200 (78.42) | 8784 (84.32) | 9610 (91.78) | <0.0001 |
| FBG (mmol/L) | 5.04±0.88 | 5.09±0.95 | 5.20±1.06 | 5.42±1.48 | <0.0001 |
| TG (mmol/L) | 0.65±0.19 | 1.02±0.23 | 1.44±0.37 | 3.22±1.93 | <0.0001 |
| TC (mmol/L) | 4.48±0.88 | 4.61±0.92 | 4.78±0.95 | 4.81±1.33 | <0.0001 |
| Heart rate (bpm) | 73.61±10.28 | 73.80±9.89 | 74.35±10.03 | 75.43±10.18 | <0.0001 |
| eGFR(mL/min/1.73m<sup>2</sup>) | 100.30±21.49 | 95.07±23.38 | 96.53±22.51 | 95.31±23.38 | <0.0001 |
| Waist circumference (cm) | 80.01±8.73 | 82.59±8.67 | 85.23±8.76 | 88.59±8.61 | <0.0001 |
| Hip circumference (cm) | 91.66±8.63 | 93.35±8.72 | 95.50±9.04 | 98.17±8.81 | <0.0001 |
| Waist-to-hip ratio | 0.88±0.08 | 0.89±0.07 | 0.89±0.07 | 0.90±0.06 | <0.0001 |
| BMI (kg/m<sup>2</sup>) | 22.77±3.11 | 23.97±3.32 | 25.05±3.54 | 26.37±3.52 | <0.0001 |
| Systolic BP (mmHg) | 117.56±15.21 | 120.59±15.50 | 122.76±15.64 | 126.91±16.46 | <0.0001 |
| Diastolic BP(mmHg) | 76.94±10.06 | 79.18±10.37 | 80.68±10.76 | 83.54±11.38 | <0.0001 |
| LDL-C (mmol/L) | 2.31±0.76 | 2.48±0.68 | 2.55±0.73 | 2.47±0.84 | <0.0001 |
| HDL-C (mmol/L) | 1.64±0.38 | 1.47±0.30 | 1.37±0.30 | 1.28±0.35 | <0.0001 |
| Hypertension, n (%) | 601 (5.73) | 741 (7.09) | 1024 (9.83) | 1690 (16.14) | <0.0001 |
| Diabetes, n (%) | 51<br>(0.49) | 74<br>(0.71) | 115<br>(1.10) | 220<br>(2.10) | <0.0001 |
| Never smoking, n<br>(%) | 6822<br>(65.08) | 6657<br>(63.67) | 6021 (57.79) | 5206<br>(49.72) | <0.0001 |
| Never drinking,<br>n (%) | 6778 (64.66) | 6739 (64.45) | 6104 (58.59) | 5445<br>(52.00) | <0.0001 |

Results reported as means ± standard deviation (SD) or percentage, unless otherwise indicated
BMI: body mass index; BP: blood pressure; eGFR: estimated glomerular filtration rate; FBG: fasting blood glucose; HDL-C: high-density lipoprotein cholesterol; LDL-C: low-density lipoprotein cholesterol; TC: total cholesterol; TG: triglyceride

### Incidence of CVD, stroke, and all-cause mortality by AIP quartile in early adulthood

During an average follow-up period of 12.65±3.59 years, we identified 1,113 incident cases of CVD, 953 cases of stroke, and 969 cases of all-cause mortality. The CVD cases comprised 174 MI, 822 IS, and 145 HS. As shown in the Kaplan–Meier curves in Figure 2, the cumulative incidence of CVD, stroke, and all-cause mortality increased progressively across ascending AIP quartiles. The cumulative incidence of CVD was 1.57% in Q1, 2.32% in Q2, 3.07% in Q3, and 3.68% in Q4. Similarly, the cumulative incidence of stroke was 1.44% in Q1, 2.03% in Q2, 2.51% in Q3, and 3.13% in Q4. The cumulative incidence of all-cause mortality was 1.80% in Q1, 2.03% in Q2, 2.51% in Q3, and 2.92% in Q4. The incidence rates per 1,000 person-years also increased with higher AIP quartiles (Table 2). For CVD, the rates were 1.27 (Q1), 1.80 (Q2), 2.41 (Q3), and 2.93 (Q4). For stroke, the rates were 1.16 (Q1), 1.57 (Q2), 1.97 (Q3), and 2.49 (Q4). For all-cause mortality, the rates were 1.44 (Q1), 1.56 (Q2), 1.95 (Q3), and 2.30 (Q4).

**Fig. 2.**
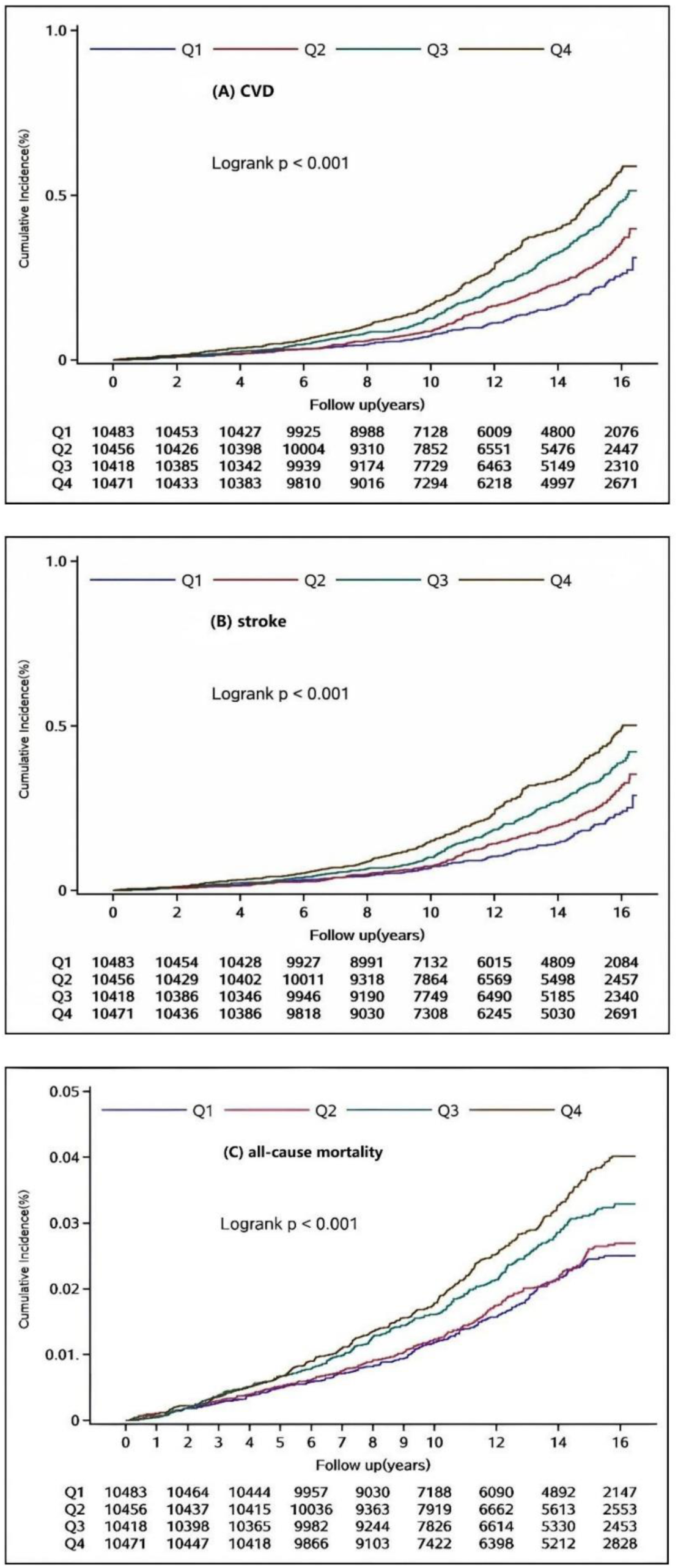
Kaplan-Meier curves for CVD, stroke and all-cause mortality according to the AIP quartiles. **A** CVD, **B** stroke, and **C** all-cause mortality. AIP: Atherogenic index of plasma; CVD: cardiovascular diseases; Q: quartile

**Table 2.**
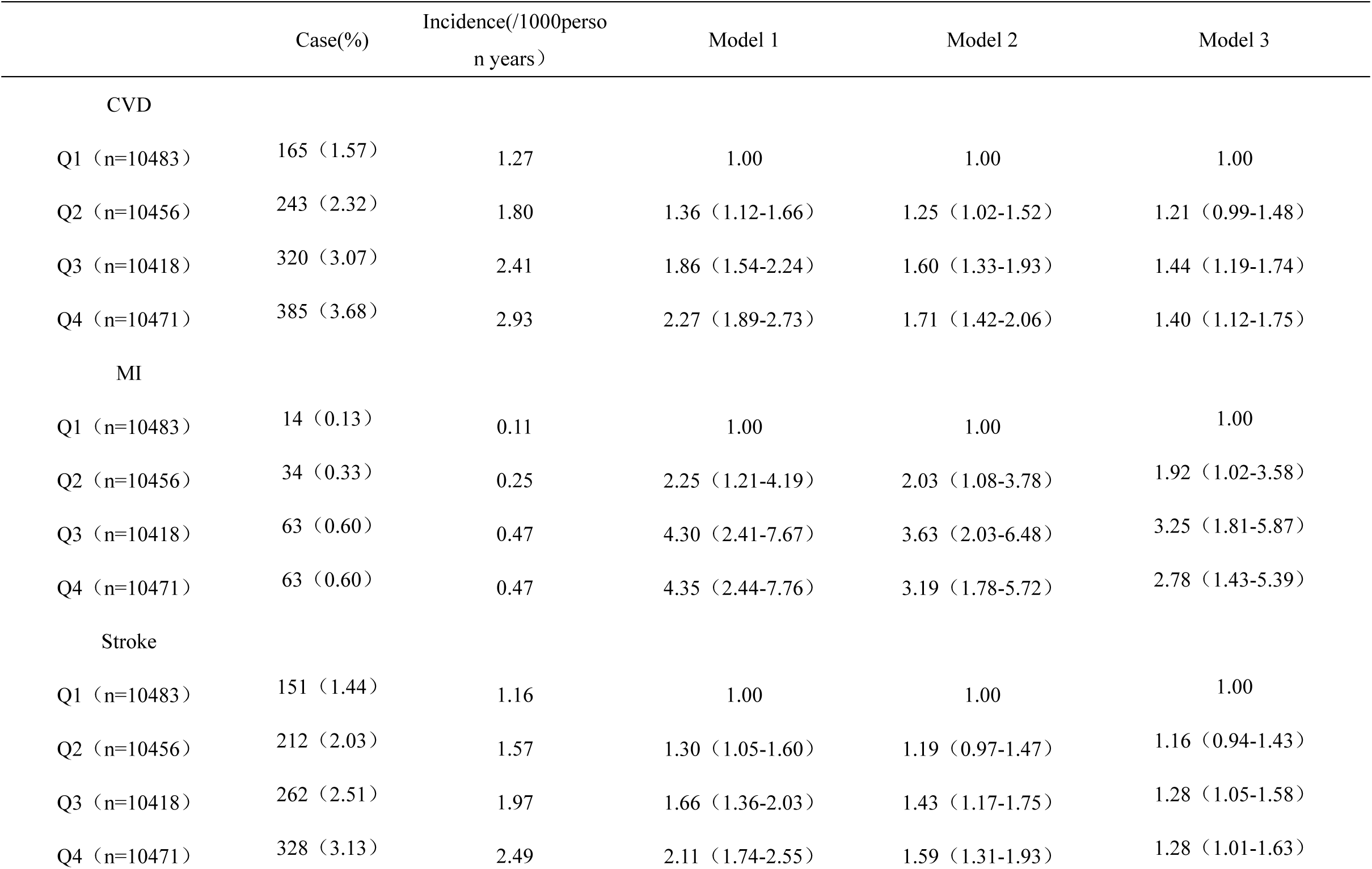
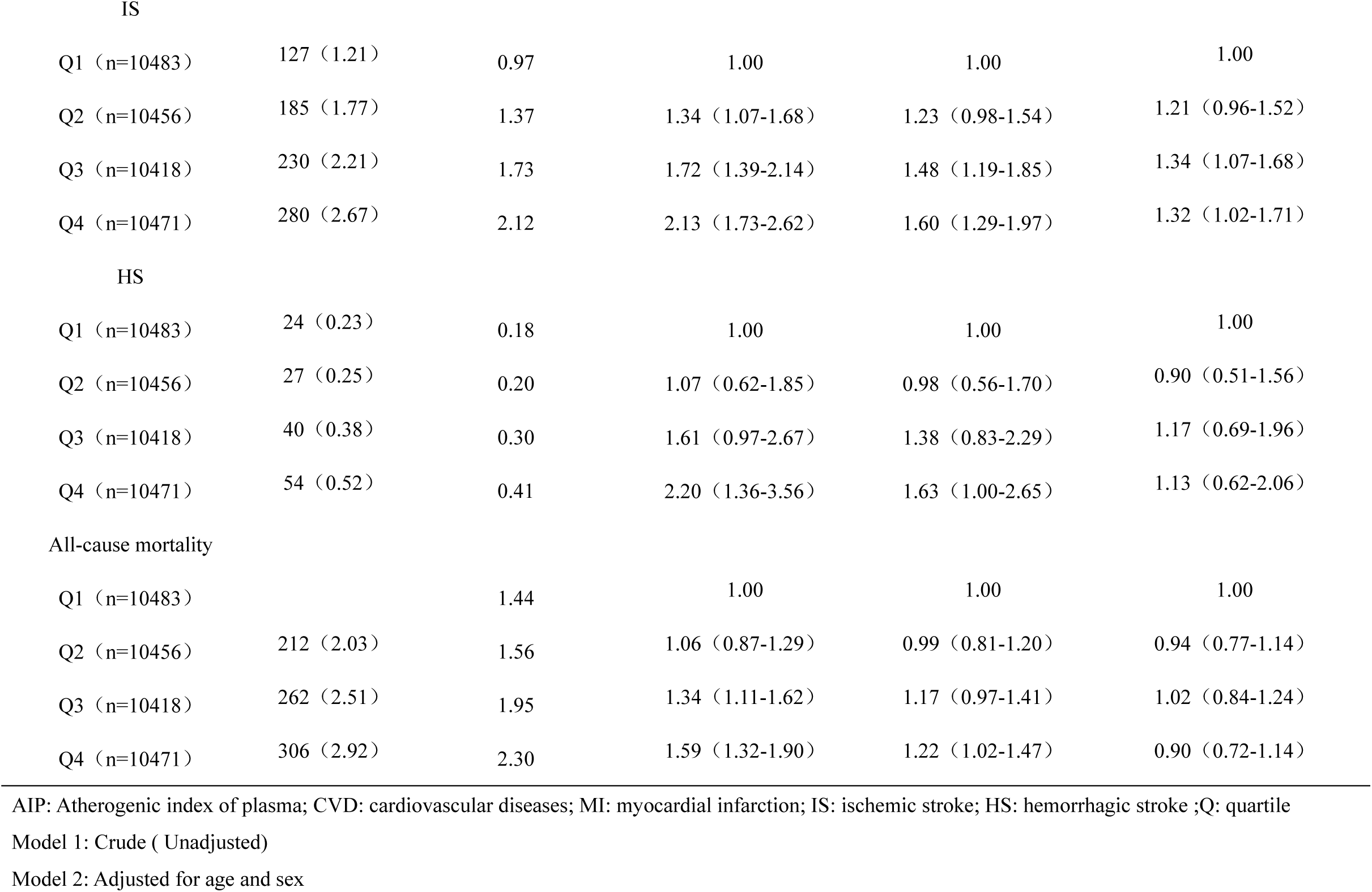
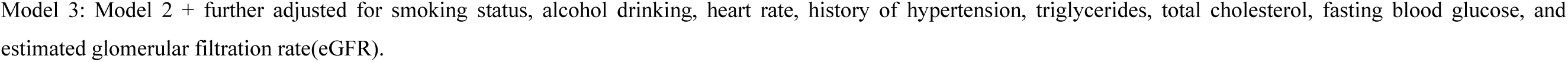
AIP Levels in Early Adulthood and CVD and All-cause mortality Risk.

### Relationship between AIP and the risks for CVD

Compared with the first quartile (Q1), the age-and sex-adjusted hazard ratios (HRs) for CVD were significantly elevated in Q2 (HR 1.25; 95% CI 1.02–1.52), Q3 (HR 1.60; 95% CI 1.33–1.93), and Q4 (HR 1.71; 95% CI 1.42–2.06). A similar increasing trend was observed for stroke: Q2: HR 1.19, 95% CI 0.97–1.47; Q3: HR 1.43, 95% CI 1.17–1.75; Q4: HR 1.59, 95% CI 1.31–1.93. For all-cause mortality, significantly increased HRs were observed only in Q4 relative to Q1: Q2: HR 0.99, 95% CI 0.81–1.20; Q3: HR 1.17, 95% CI 0.97–1.41; Q4: HR 1.22, 95% CI 1.02–1.47. Following multivariate adjustment for age, gender, smoking status, alcohol consumption, heart rate, hypertension history, TG, TC, FBG, and eGFR, a significant and graded increase in risk persisted for both CVD and stroke across higher AIP quartiles. The fully adjusted HRs for CVD were as follows: Q2: 1.21 (95% CI 0.99–1.48); Q3: 1.44 (95% CI 1.19–1.74); Q4: 1.40 (95% CI 1.12–1.75). Similarly, the adjusted HRs for stroke were: Q2: 1.16 (95% CI 0.94–1.43); Q3: 1.28 (95% CI 1.05–1.58); Q4: 1.28 (95% CI 1.01–1.63) (Table 2). In contrast, no significant association was observed between AIP quartiles and all-cause mortality after full adjustment. The multivariate-adjusted hazard ratios for CVD, stroke, and all-cause mortality across AIP quartiles are summarized in Table 2.

Sensitivity analyses, which excluded participants who developed CVD within the first 2 years of follow-up, those with cancer at baseline, or those using antihypertensive, hypoglycemic, or lipid-lowering medications (Supplemental Table S1), yielded consistent results with the primary analysis. In subtype analyses stratified by age, sex, fasting blood glucose (FBG) level, and hypertension status (Supplemental Table S2), higher AIP quartiles remained significantly associated with an increased risk of myocardial infarction (MI) and ischemic stroke (IS), but not hemorrhagic stroke (HS).

Stratified analyses indicated that BMI significantly modified the association between AIP and all-cause mortality (P for interaction = 0.02). In contrast, no significant interaction impact was observed for the other stratification variables (all *P* for interaction > 0.05; Supplemental Tables S2–S4).

## Discussion

This large prospective cohort study establishes a significant and independent association between elevated AIP and an increased risk of incident CVD and stroke in adults aged 18–44 years. To our knowledge, this study identified AIP as an early metabolic predictor of cardiovascular diseases in young adults, with a graded dose-response relationship. Therefore, AIP may be a potential tool for identifying cardiovascular diseases risk in early adulthood.

Our findings extend previous research on the association between AIP and cardiovascular risk, previously established in diabetic patients^14^, nondiabetic hypertensive older adults^15^, healthy adults^29^, and the general population^16,17^, to young population. Adjusting for conventional risk factors, lipids, and renal function, the robust and graded increase of CVD and stroke risk observed across AIP quartiles in this demographic persisted, highlighting the value of AIP in detecting atherogenic dyslipidemia at an early stage. Notably, elevated AIP in early adulthood was associated with a higher risk of subsequent CVD and stroke. The underlying pathological mechanism of this association is atherosclerosis, because most CVD events usually begin to progress asymptotically years before the clinical onset^30,31^. Therefore, AIP measured in young adulthood may reflect the cumulative burden and progression of subclinical atherosclerosis, supporting it as a reliable marker of atherosclerotic development and a practical tool for early CVD risk stratification. Specifically, participants in the highest quartile had a 40% increased risk of CVD compared to those in the lowest quartile. A similar trend was observed for stroke. Furthermore, subtype analyses revealed consistent associations between elevated AIP and ischemic events (MI and IS), but not with HS, further supporting a pathophysiology rooted in atherosclerotic progression.

This association is biologically plausible, which is supported by the established correlation between AIP and atherogenic small dense LDL (sdLDL) particles^32^. Functionally, AIP reflects lipoprotein particle size and the balance between proatherogenic and antiatherogenic lipoproteins, making it a clinically useful surrogate marker for sdLDL^33^and a potent predictor of major adverse cardiovascular events^12,15^. Moreover, the association of AIP with carotid intima-media thickness^34^, arterial stiffness^35^, and coronary artery atherosclerosis ^36^ further strengthens its pathophysiological correlation, and confirms its role as a reliable predictor of atherosclerosis progression. No significant association AIP with all-cause mortality may be attributable to the relatively young cohort and limited follow-up duration; However, the modifying effect of BMI on this relationship indicates that metabolic status influences long-term outcomes and merits further investigation.

Our study possesses notable strengths. We demonstrated a significant association between AIP in early adulthood and subsequent risks of CVD and stroke by adjusting multiple confounding variables and conducting subgroup analyses. Therefore, early-life AIP assessment may help identify individuals with elevated cardiovascular risk before the clinical onset of disease. However, this study also has several limitations. First, our research only includes Chinese people, which may limit the generalizability of our findings to other ethnic groups. Second, the number of outcome events may have resulted in insufficient statistical power. However, our study is based on the young population without CVD, who have relatively fewer events. Finally, the parameters used for AIP calculation were measured solely at baseline, and how the changing trend of AIP over time influences future risk of CVD warrants further investigation.

## Conclusions

In conclusion, an elevated AIP increased the risk of CVD and stroke in young adults. AIP may be a biomarker for identifying young adults at a high risk of CVD. Further prospective studies are needed to investigate the impact of AIP on cardiovascular events.

## Abbreviations

AIP: Atherogenic index of plasma
CVD: Cardiovascular diseases
MI: Myocardial infarction
IS: Ischemic stroke
HS: Hemorrhagic stroke
TG: Triglyceride
HDL-C: High-density lipoprotein cholesterol
LDL-C: Low-density lipoprotein cholesterol
sdLDL: small, dense LDL
TC: Total cholesterol
BP: Blood pressure
BMI: Body mass index
FBG: Fasting blood glucose
CKD-EPI: Chronic Kidney Disease Epidemiology Collaboration
eGFR: Estimated glomerular filtration rate

## Data Availability

All data generated or analysed during this study are included in this published article (and its supplementary information files).

## ACKNOWLEDGMENTS

The authors thank all the survey teams of the Kailuan study group for their contribution and the study participants who contributed their information.

## Author contributions

H.X. and S.W. designed the study. N.Y. and M.H. conducted the data analyses. M.H. drafted the manuscript. N.Y., C.W. and Z.F. critically revised the manuscript for important intellectual content. All authors have approved the final version of the manuscript. S.W., and H.X. are the guarantors of this work and, as such, had full access to all of the data in the study and take responsibility for the integrity of the data and the accuracy of the data analysis. The corresponding author attests that all listed authors meet the authorship criteria and that no others meeting the criteria have been omitted. All authors read and approved the final manuscript.

## Data availability

No datasets were generated or analysed during the current study.

## Ethics approval and consent to participate

This study was approved by the Institutional Review Board of Kailuan General Hospital (approval number: 2006-5). We obtained written informed consent from all participants.

## Consent for publication

Not applicable.

---

**Supplemental Table S1.**
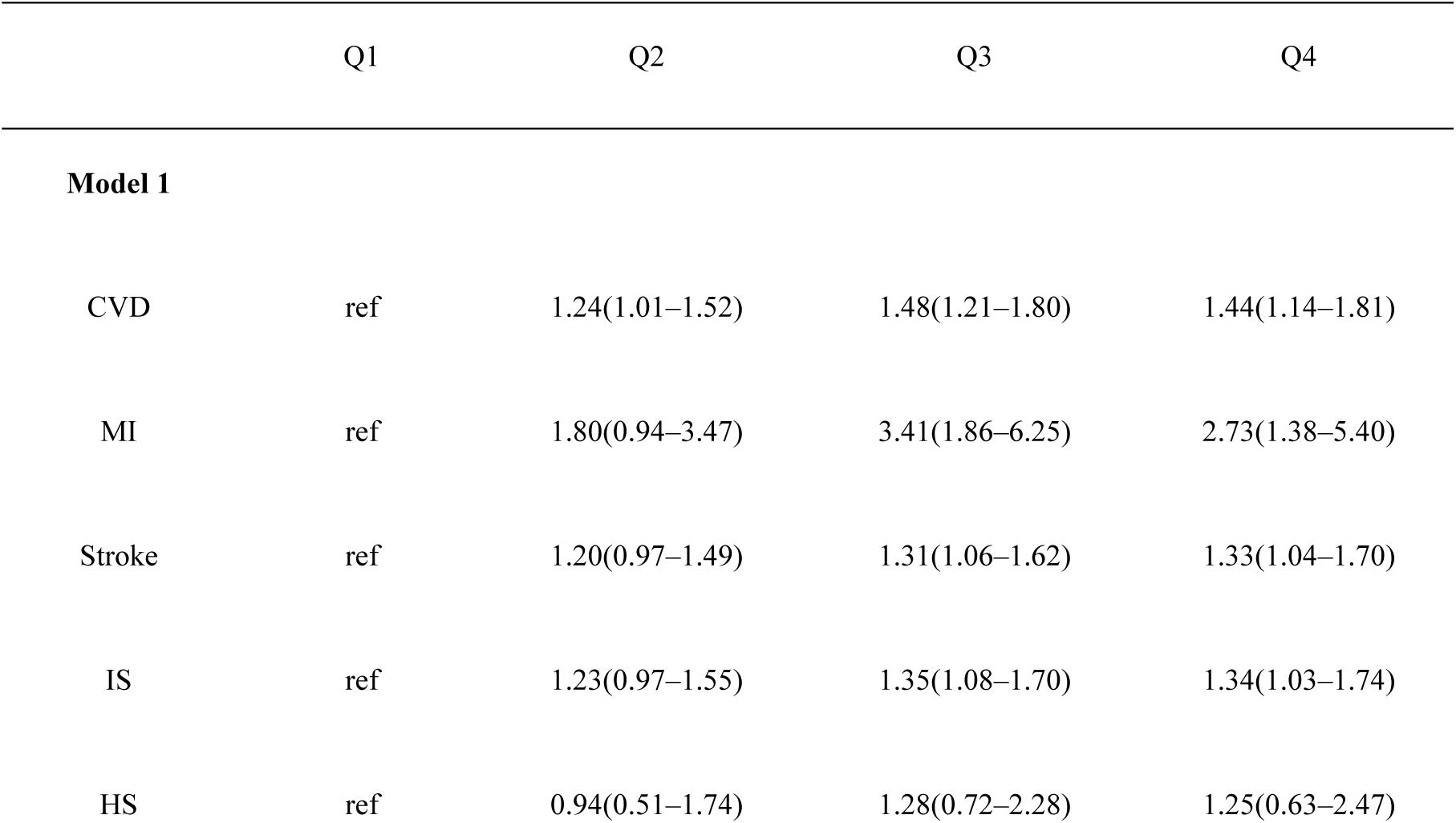
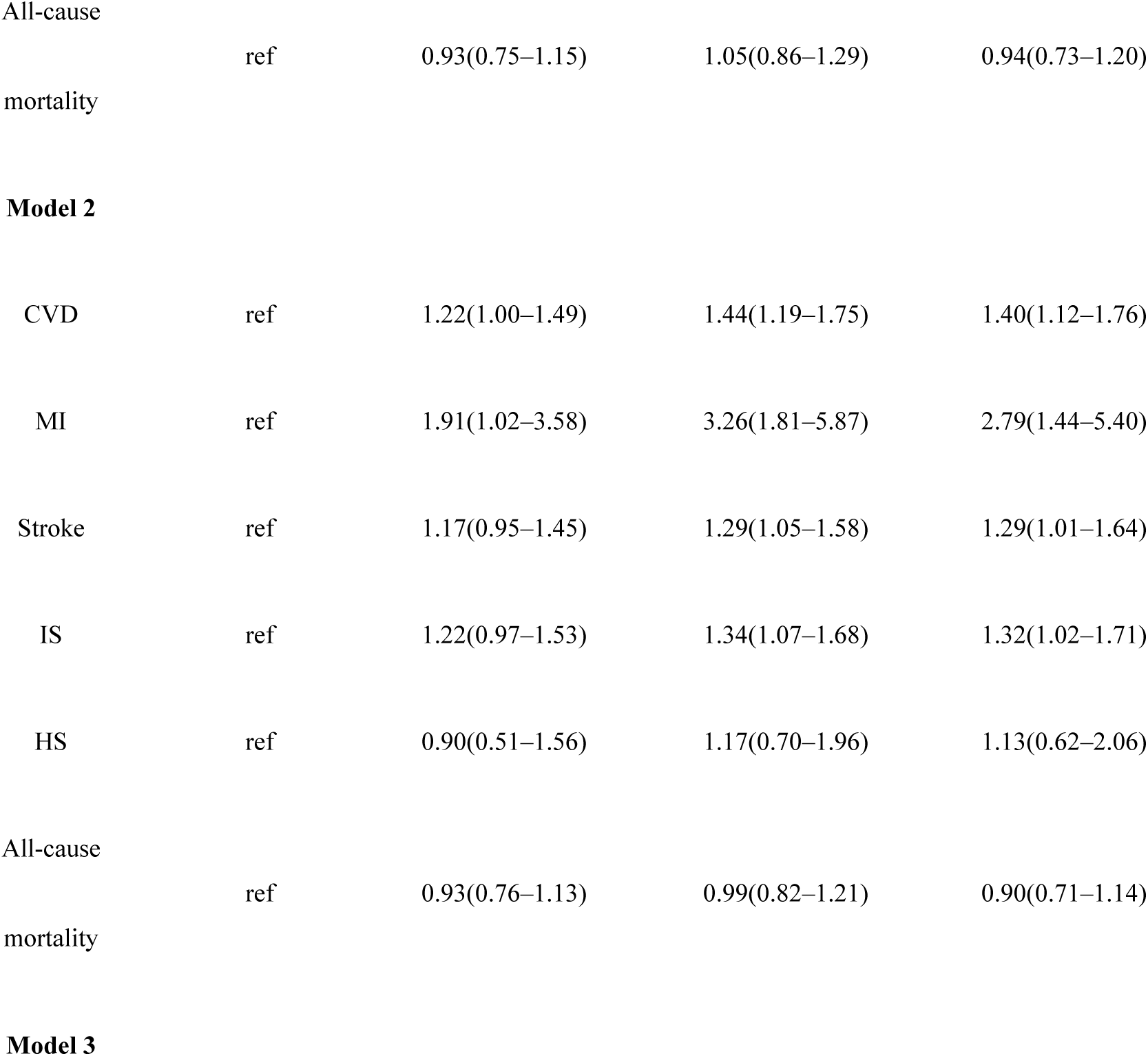
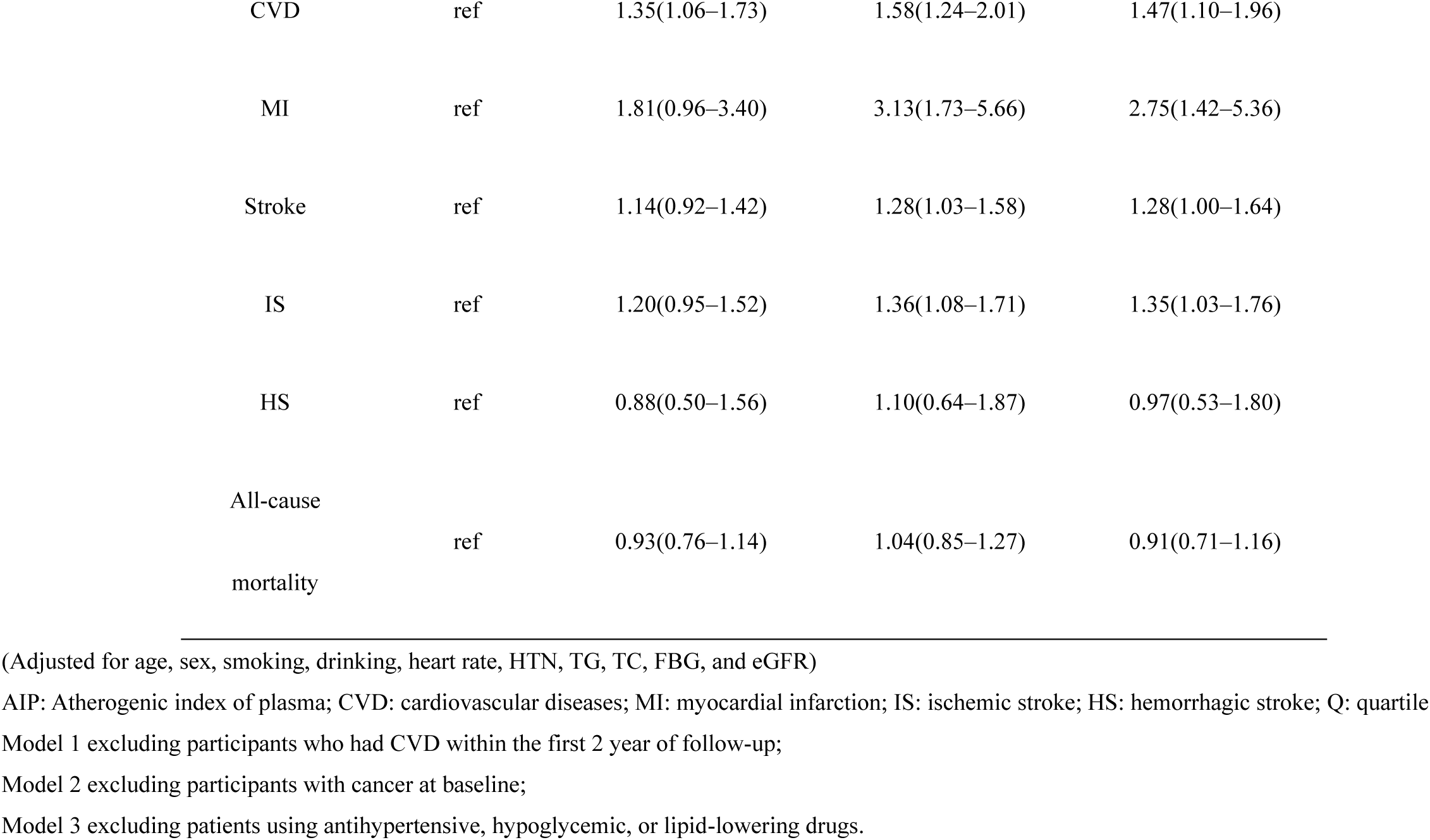
Sensitivity analysis(AIP Levels in Early Adulthood and CVD and All-cause mortality Risk)

**Supplemental Table S2.**
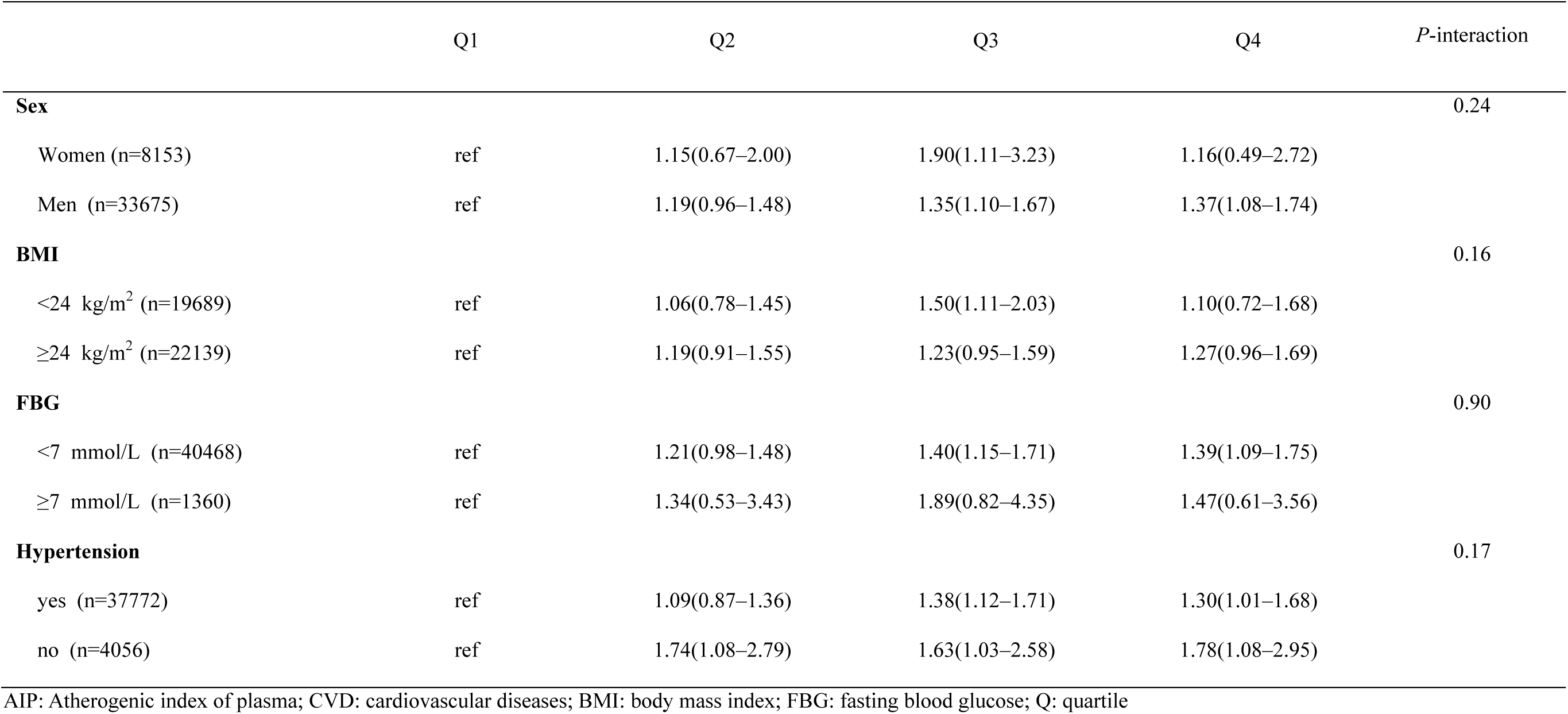
Stratified analyses for the association of AIP with CVD.

|  | Q1 | Q2 | Q3 | Q4 | *P*-interaction |
| --- | --- | --- | --- | --- | --- |
| **Sex** |  |  |  |  | 0.24 |
| Women (n=8153) | ref | 1.15(0.67–2.00) | 1.90(1.11–3.23) | 1.16(0.49–2.72) |  |
| Men (n=33675) | ref | 1.19(0.96–1.48) | 1.35(1.10–1.67) | 1.37(1.08–1.74) |  |
| **BMI** |  |  |  |  | 0.16 |
| <24 kg/m<sup>2</sup> (n=19689) | ref | 1.06(0.78–1.45) | 1.50(1.11–2.03) | 1.10(0.72–1.68) |  |
| ≥24 kg/m<sup>2</sup> (n=22139) | ref | 1.19(0.91–1.55) | 1.23(0.95–1.59) | 1.27(0.96–1.69) |  |
| **FBG** |  |  |  |  | 0.90 |
| <7 mmol/L (n=40468) | ref | 1.21(0.98–1.48) | 1.40(1.15–1.71) | 1.39(1.09–1.75) |  |
| ≥7 mmol/L (n=1360) | ref | 1.34(0.53–3.43) | 1.89(0.82–4.35) | 1.47(0.61–3.56) |  |
| **Hypertension** |  |  |  |  | 0.17 |
| yes (n=37772) | ref | 1.09(0.87–1.36) | 1.38(1.12–1.71) | 1.30(1.01–1.68) |  |
| no (n=4056) | ref | 1.74(1.08–2.79) | 1.63(1.03–2.58) | 1.78(1.08–2.95) |  |

AIP: Atherogenic index of plasma; CVD: cardiovascular diseases; BMI: body mass index; FBG: fasting blood glucose; Q: quartile

**Supplemental Table S3.**
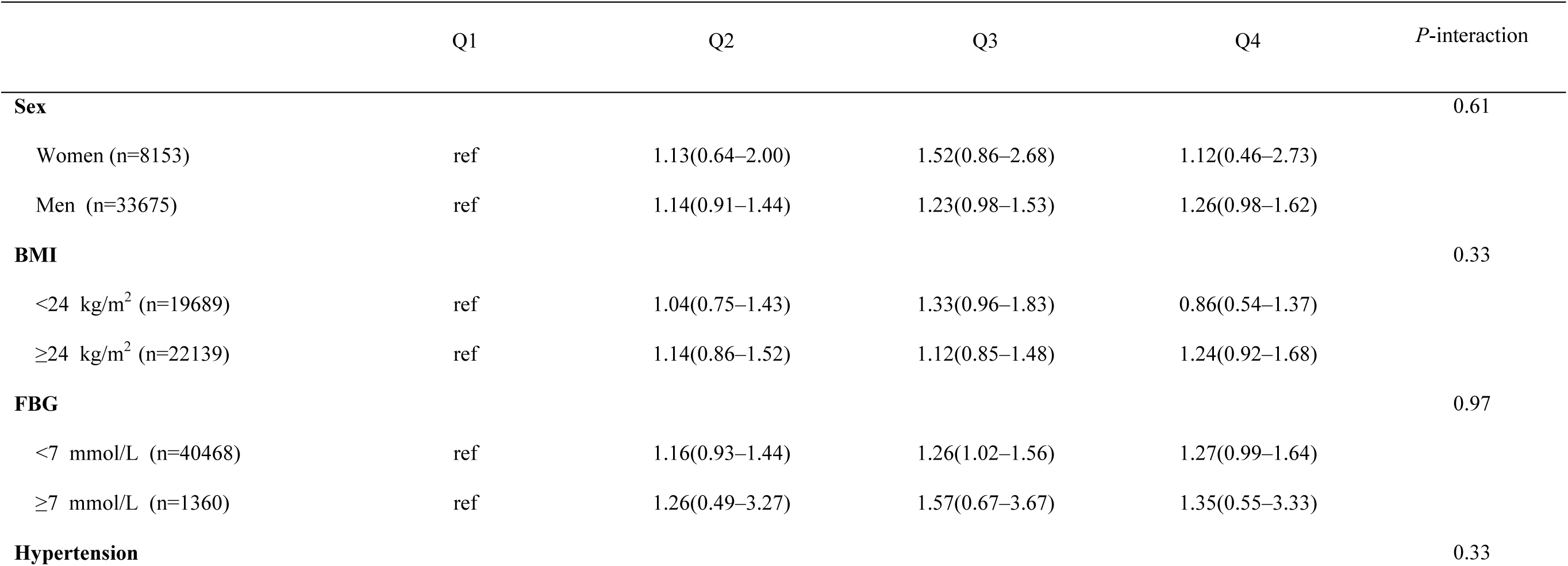
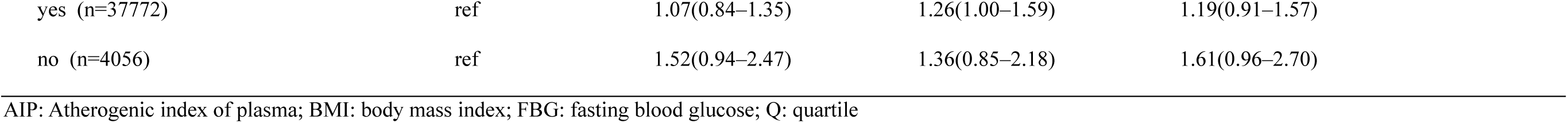
Stratified analyses for the association of AIP with stroke.

|  | Q1 | Q2 | Q3 | Q4 | *P*-interaction |
| --- | --- | --- | --- | --- | --- |
| **Sex** |  |  |  |  | 0.61 |
| Women (n=8153) | ref | 1.13(0.64–2.00) | 1.52(0.86–2.68) | 1.12(0.46–2.73) |  |
| Men (n=33675) | ref | 1.14(0.91–1.44) | 1.23(0.98–1.53) | 1.26(0.98–1.62) |  |
| **BMI** |  |  |  |  | 0.33 |
| <24 kg/m<sup>2</sup> (n=19689) | ref | 1.04(0.75–1.43) | 1.33(0.96–1.83) | 0.86(0.54–1.37) |  |
| ≥24 kg/m<sup>2</sup> (n=22139) | ref | 1.14(0.86–1.52) | 1.12(0.85–1.48) | 1.24(0.92–1.68) |  |
| **FBG** |  |  |  |  | 0.97 |
| <7 mmol/L (n=40468) | ref | 1.16(0.93–1.44) | 1.26(1.02–1.56) | 1.27(0.99–1.64) |  |
| ≥7 mmol/L (n=1360) | ref | 1.26(0.49–3.27) | 1.57(0.67–3.67) | 1.35(0.55–3.33) |  |
| **Hypertension** |  |  |  |  | 0.33 |

|  |  |  |  |  |
| --- | --- | --- | --- | --- |
| yes (n=37772) | ref | 1.07(0.84–1.35) | 1.26(1.00–1.59) | 1.19(0.91–1.57) |
| no (n=4056) | ref | 1.52(0.94–2.47) | 1.36(0.85–2.18) | 1.61(0.96–2.70) |

AIP: Atherogenic index of plasma; BMI: body mass index; FBG: fasting blood glucose; Q: quartile

**Supplemental Table S4.** Stratified analyses for the association of AIP with death.

|  | Q1 | Q2 | Q3 | Q4 | *P*-interaction |
| --- | --- | --- | --- | --- | --- |
| **Sex** |  |  |  |  | 0.12 |
| Women (n=8153) | ref | 1.18(0.68–2.02) | 1.83(1.03–3.26) | 2.65(1.12–6.26) |  |
| Men (n=33675) | ref | 0.90(0.72–1.11) | 0.95(0.77–1.16) | 0.82(0.64–1.04) |  |
| **BMI** |  |  |  |  | 0.02 |
| <24 kg/m<sup>2</sup> (n=19689) | ref | 1.09(0.82–1.44) | 1.28(0.96–1.70) | 1.19(0.79–1.78) |  |
| ≥24 kg/m<sup>2</sup> (n=22139) | ref | 0.77(0.58–1.02) | 0.83(0.63–1.08) | 0.74(0.55–1.00) |  |
| **FBG** |  |  |  |  | 0.72 |
| <7 mmol/L (n=40468) | ref | 0.92(0.75–1.13) | 1.02(0.84–1.25) | 0.87(0.68–1.12) |  |
| ≥7 mmol/L (n=1360) | ref | 1.04(0.49–2.20) | 1.02(0.50–2.07) | 1.08(0.50–2.33) |  |
| **Hypertension** |  |  |  |  | 0.31 |
| yes (n=37772) | ref | 0.92(0.74–1.16) | 1.11(0.89–1.38) | 0.97(0.74–1.28) |  |
| no (n=4056) | ref | 1.01(0.66–1.56) | 0.81(0.53–1.22) | 0.77(0.49–1.21) |  |

AIP: Atherogenic index of plasma; BMI: body mass index; FBG: fasting blood glucose; Q: quartile

